# State-wise estimates of current hospital beds, intensive care unit (ICU) beds and ventilators in India: Are we prepared for a surge in COVID-19 hospitalizations?

**DOI:** 10.1101/2020.06.16.20132787

**Authors:** Geetanjali Kapoor, Stephanie Hauck, Aditi Sriram, Jyoti Joshi, Emily Schueller, Isabel Frost, Ruchita Balasubramanian, Ramanan Laxminarayan, Arindam Nandi

**Affiliations:** Center for Disease Dynamics, Economics & Policy, New Delhi, India; Center for Disease Dynamics, Economics & Policy, Washington DC, USA; Amity University, India; Imperial College London, UK; Princeton Environmental Institute, Princeton University, Princeton, USA

## Abstract

**Background:** The rapid spread of COVID-19 globally has prompted policymakers to evaluate the capacity of health care infrastructure in their communities. Many hard-hit localities have witnessed a large influx of severe cases that strained existing hospitals. As COVID-19 spreads in India, it is essential to evaluate the country’s capacity to treat severe cases.

**Methods:** We combined data on public and private sector hospitals in India to produce state level estimates of hospital beds, ICU beds, and mechanical ventilators. Based on the number of public sector hospitals from the 2019 National Health Profile (NHP) of India and the relative proportions of public and private health care facilities from the National Sample Survey (NSS) 75^th^ round (2017-2018), we estimated capacity in each Indian state and union territory (UT). We assumed that 5% of all hospital beds were ICU beds and that 50% of ICU beds were equipped with ventilators.

**Results:** We estimated that India has approximately 1.9 million hospital beds, 95,000 ICU beds and 48,000 ventilators. Nationally, resources are concentrated in the private sector (hospital beds: 1,185,242 private vs 713,986 public; ICU beds: 59,262 private vs 35,699 public; ventilators: 29,631 private vs. 17,850 public). Our findings suggest substantial variation in available resources across states and UTs.

**Conclusion:** Some projections shave suggested a potential need for approximately 270,000 ICU beds in an optimistic scenario, over 2.8 times the estimated number of total available ICU beds in India. Additional resources will likely be required to accommodate patients with severe COVID-19 infections in India.

## Introduction

COVID-19 was declared a global pandemic by the World Health Organization on 11th March 2020. This novel disease has been characterized in a variety of country contexts by a low number of initial cases followed by a sudden, “bomb-like” explosion in infections[1]. Despite widespread lockdowns to reduce transmissions, many countries’ healthcare systems have been overwhelmed by the demand for hospitals, beds, and supportive equipment needed to treat severe cases of the disease. India has already reported over 20,000 COVID-19 cases as of 22nd April 2020[2]. Although this number indicates low prevalence of COVID-19 in India’s total population, low testing rates of 0.0003 per capita (as of 22^nd^ April 2020)[3] may obscure the full extent of COVID-19 infections in the country. Public health experts have warned that sharp increases in community transmission of COVID-19 in the upcoming months will lead to a large influx of critically ill patients and subsequent demand for ICU care and mechanical ventilation.

Indian health protocols [4] dictate that anyone who tests positive for COVID-19 is placed in an isolation ward or, for critical patients, in an ICU. In addition, all suspected cases of COVID-19 are referred to government hospitals for testing[3]. The number of public hospital beds in India has been estimated at 0.55 per 1000 people, representing 12 states comprising 70% of India’s total population[5]. Of available beds, fewer than 5% of beds in public hospitals are estimated to be ICU capable.[6] Although current COVID-19 cases are being managed primarily through these public facilities, a large increase in severe infections will likely necessitate utilization of private sector resources, as well. However, the cost of private healthcare is a barrier for many Indians[7][5]. In rural India, where approximately 80% of the population live, ICU care is poor or absent at the district level, as is access to speciality care such as ventilation[6].

We estimated the number of hospital beds, Intensive Care Unit (ICU) beds, and ventilators across states and Union Territories (UTs) for both public and private facilities. These estimates, evaluated alongside projected future COVID-19 infections, may equip policy makers and healthcare professionals to tackle the COVID-19 pandemic.

## Methods

We obtained estimates of government hospital beds at national and state levels from the 2019 National Health Profile (NHP) of India.[8] These data were not available for the private sector, but we obtained the proportions of public (including charitable or NGO) and private health care facilities from the National Sample Survey (NSS) 75^th^ round (2017-2018)[9]. NSS 75^th^, a nationally representative health survey of 113,823 households in India, collected self-reported information on hospitalizations during 365 days preceding the survey. Based on these proportions and the number of public hospital beds from the NHP, we estimated the number of private hospital beds in each state. The state level estimates were then aggregated to produce a national estimate of private hospital beds.

It is estimated that ICU beds constitute 5-8% of total beds available in large public teaching hospitals. We assumed that 5% of all hospital beds in both public and private facilities were ICU beds, and that 50% of all ICU beds were equipped with ventilators.

## Results

We estimated that India has approximately 1.9 million hospital beds, 95,000 ICU beds and 48,000 ventilators. Nationally, hospital beds are concentrated in the private sector (hospital beds: 1,185,242 private vs 713,986 public). ICU beds and ventilators follow a similar trend (ICU beds: 59,262 private vs 35,699 public; ventilators: 29,631 private: 17,850 public).

Most hospital beds and ventilators in India are concentrated in seven States - Uttar Pradesh (14.8%), Karnataka (13.8%), Maharashtra (12.2%), Tamil Nadu (8.1%), West Bengal (5.9%), Telangana (5.2%) and Kerala (5.2%) (Figure 1 and 3, Supplemental Tables 1 and 2). Our findings suggest substantial variation in available resources across states and UTs. Small UTs such as Daman and Diu, Chandigarh, and Puducherry have among the most estimated hospital beds per capita, with 906, 510, and 375 beds per 100,000 population, respectively, while Karnataka also has a relatively large capacity at 392 beds per 100,000 (Supplemental Tables 3 and 4). Conversely, Bihar, Odisha, and Chhattisgarh are estimated to have the fewest beds per population, with 26, 55, and 56 beds per 100,000, respectively. Number of ICU beds and ventilators per 100,000 population follows similar trends (Supplemental Tables 3 and 4).

**Figure 1:**
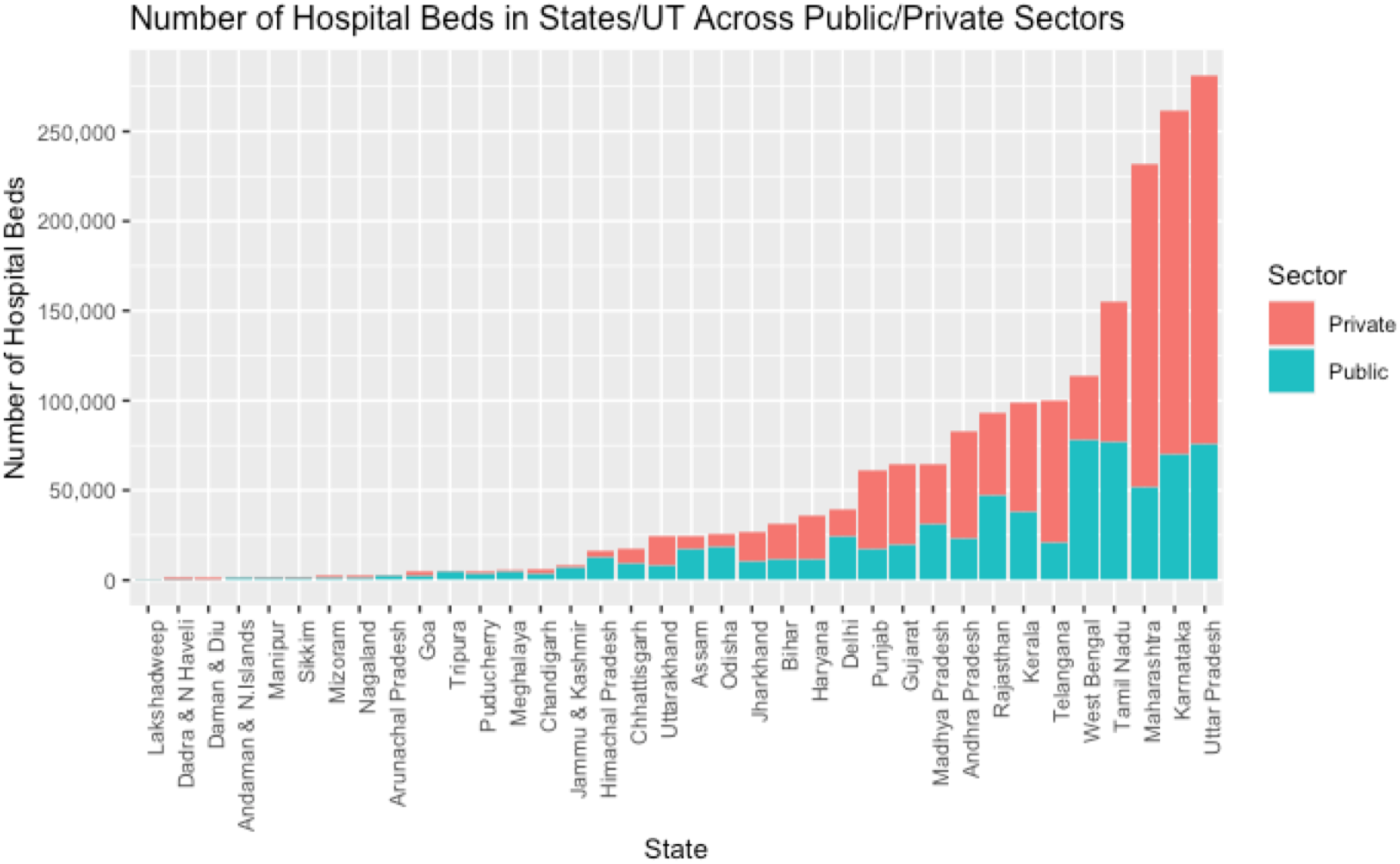
Number of hospital beds in both public and private sector, where private sector numbers are estimated values. States/UTs have been arranged in increasing order of total number of hospital beds. Height of column represents total number of hospital beds in that State.

**Figure 2:**
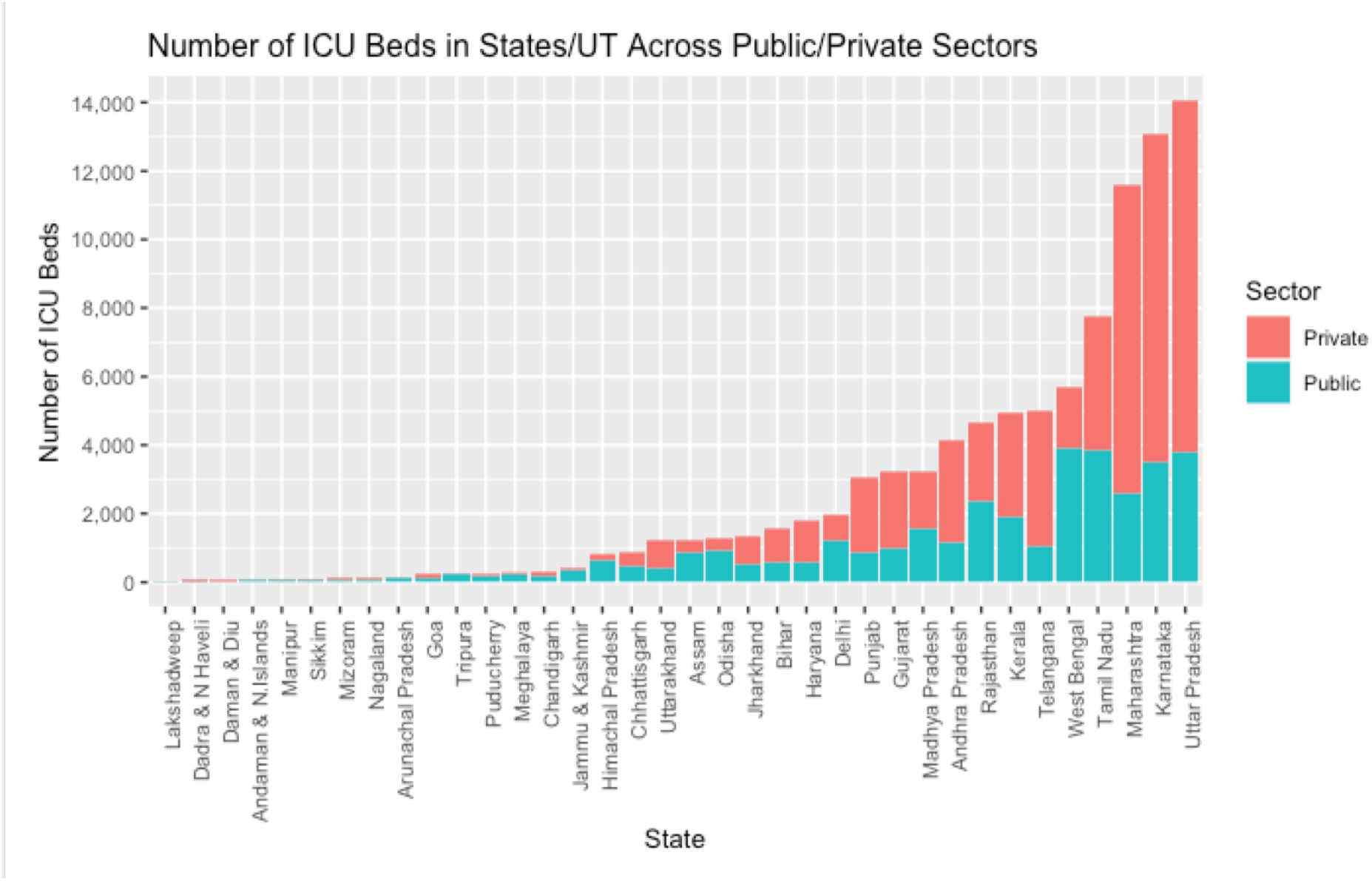
Numbers of ICU beds (both in public and private sectors), are estimated values. States/UTs have been arranged in increasing order of total number of ICU beds. Height of column represents total number of ICU beds in that State.

**Figure 3:**
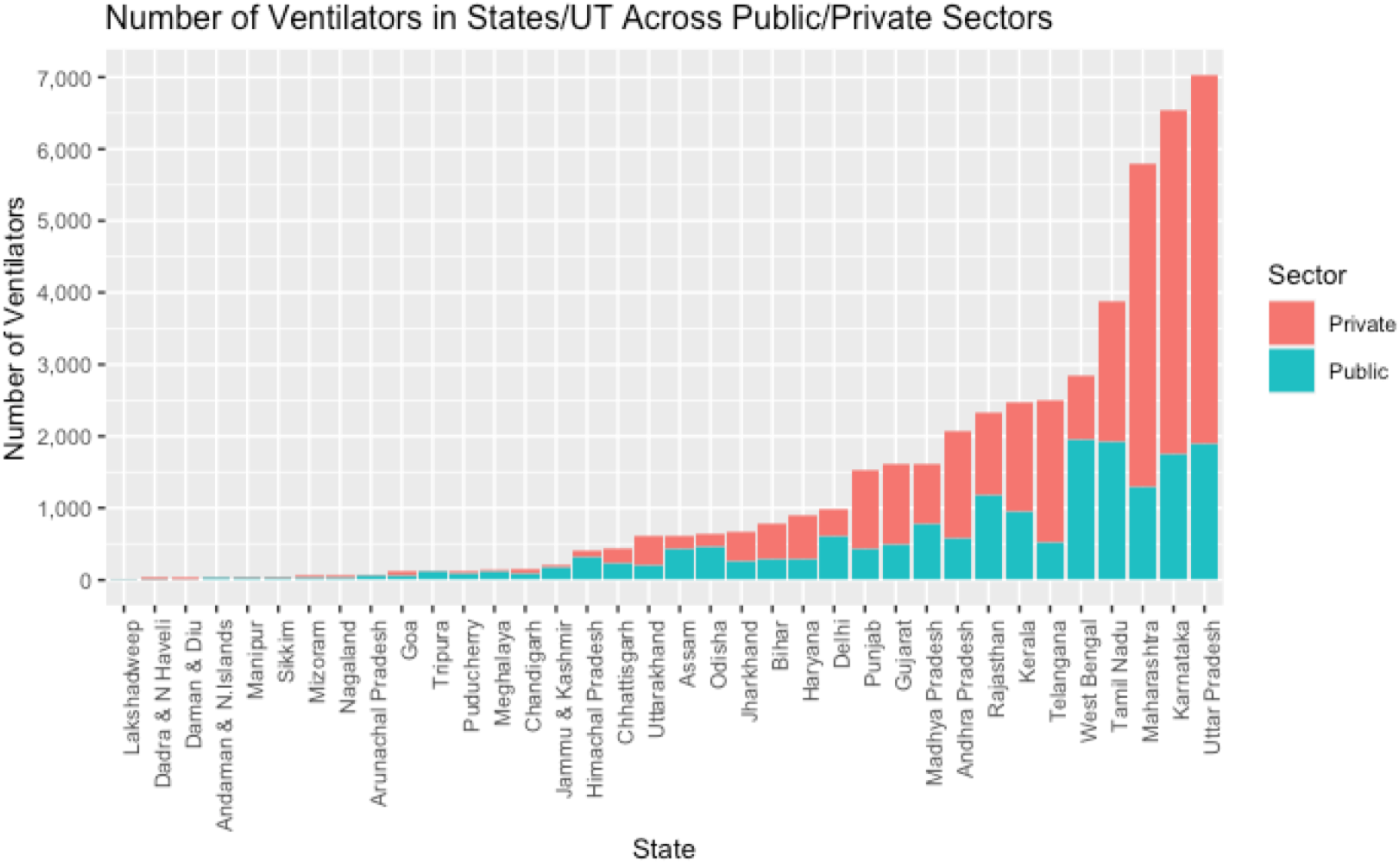
Numbers of ventilators (both in public and private sectors), are estimated values. States/UTs have been arranged in increasing order of total number of ventilators. Height of column represents total number of ventilators in that State.

## Discussion

Depending on the effectiveness of interventions, severe COVID-19 infections in India that require hospitalization are projected to peak between 673,822-2.8 million before July 2020.[10] Due to the critical nature of severe COVID-19 infection, including severe pneumonia and/or multiple systems failure, we estimate that 40-50% of hospitalized infections from COVID-19 will require ICU beds. This implies a potential need for approximately 270,000 ICU beds in the most optimistic scenario, over 2.8 times the estimated number of total available ICU beds in India, and a large portion of these will further require ventilators.

Some of the hardest hit and richest nations have been overwhelmed by the surge in demand for hospital beds, ventilators and other essentials of acute care. The World Health Organizations does not have a global recommendation for the number of hospital beds per 1000 population, leaving policymakers to make educated guesses based on currently known data on the disease.[11] For reference, the United States has 2.77 beds per 1000 people and 29.4 ICU beds per 100,000, with 18.8 ventilators per 100,000 people. Italy has 3.17 beds per 1000 and 12.5 ICU beds per 100,000 with 8.3 ventilators per 100,000 people.[12] China, which is the largest middle income country, has 4.05 beds per 1000 people and 3.6 ICU beds/100,000 and unknown numbers of ventilators.[12]

It is expected that elderly COVID-19 patients and those with pre-existing conditions will require all existing hospital resources, as they will be more severely affected by COVID-19 than younger segments of the population. These patients will likely require critical care and potentially require supportive/mechanical ventilation. Historic ICU utilization rate trends across India suggest that during monsoon seasons (which differ between the north and south of the country), 70-80% of admitted patients have an infectious disease (malaria, dengue), with the remaining patients affected with metabolic diseases such as diabetes or severe cardiac conditions[6]. Concurrently, 30-40% of long-term ICU patients are elderly (>80 years old) and are at an extreme risk for nosocomial infection within ICU treatment wards, including COVID-19[6].

Public mass testing could provide a more complete picture of the spread of COVID-19 in India but has yet to be rolled out due to limitations regarding testing facilities, equipment, trained personnel, availability of effective tests and high associated costs. Per existing guidelines, only a doctor can recommend a patient for a COVID-19 test after a suspected case presents at a facility or through the home care network, although some private labs began to offer tests to private citizens for INR 3500-4500 ($46-59.15 USD) as of April 20, 2020.[13] At the time of writing this report, 235 government and 86 private laboratories are authorized to test for COVID-19 in India.[3] This expansion in access to testing will provide useful data to inform the Indian health response to COVID-19; additionally, mass testing, facilitated by the pooling of multiple tests to minimize resource consumption[14], is necessary to monitor COVID-19 spread across India and identify hotspots of infection.

Health policy is generally relegated to Indian state governments rather than the national government; therefore, our estimates seek to fill the gap in existing data sources to estimate the total resources available for critically ill patients in each Indian state and union territory. This analysis suggests that the availability of total beds, ICU beds, and ventilators in India is insufficient to handle a large influx of severely ill COVID-19 patients in addition to their utilization for other acute illnesses. Estimates on expected demand for critical care services will enable policymakers to prepare health facilities for a sharp rise in severe COVID-19 cases.

## Data Availability

Data are publicly available from published studies and reports as cited in the manuscript.

## Supplemental Tables

**Supplemental Table 1:**
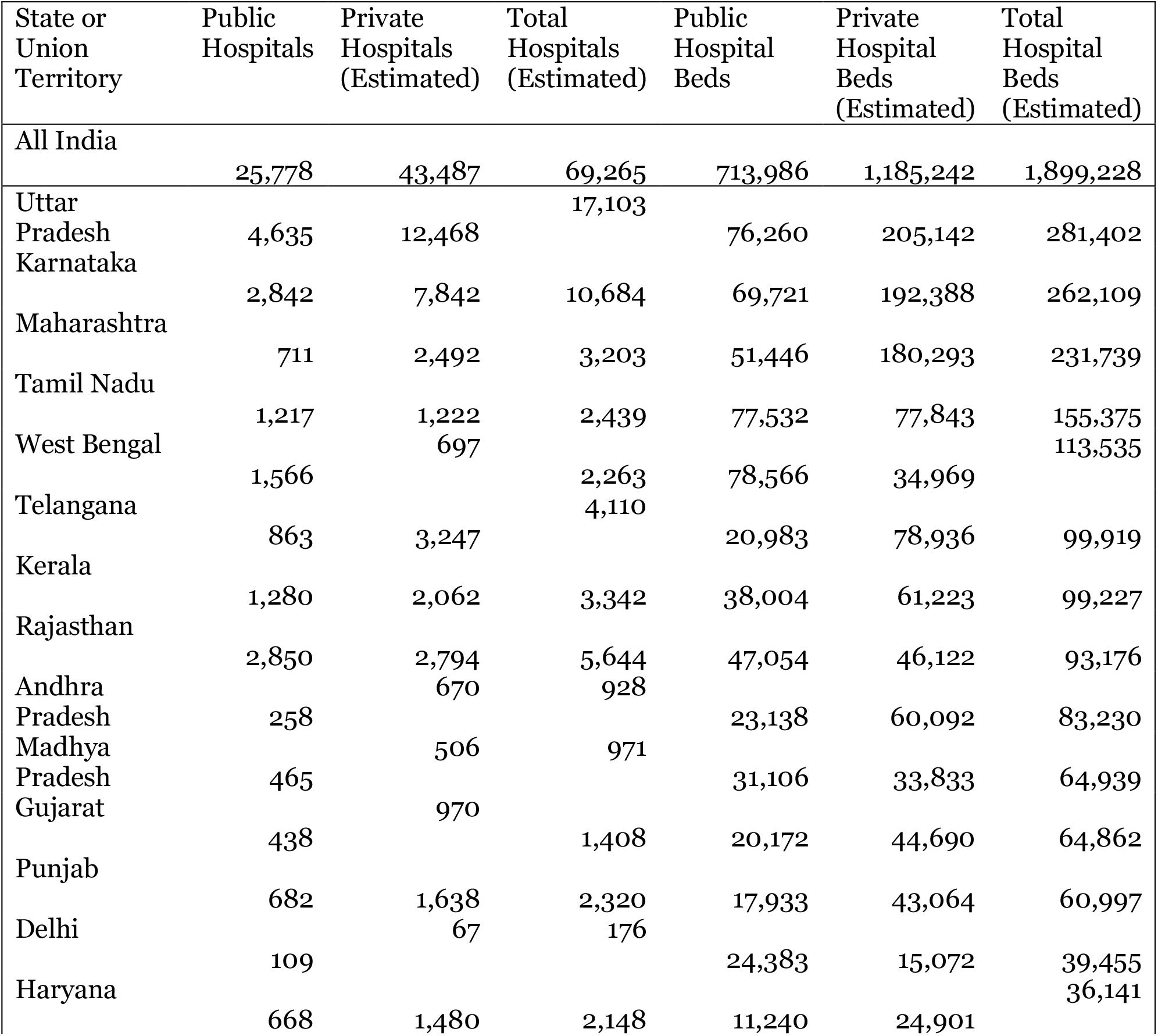
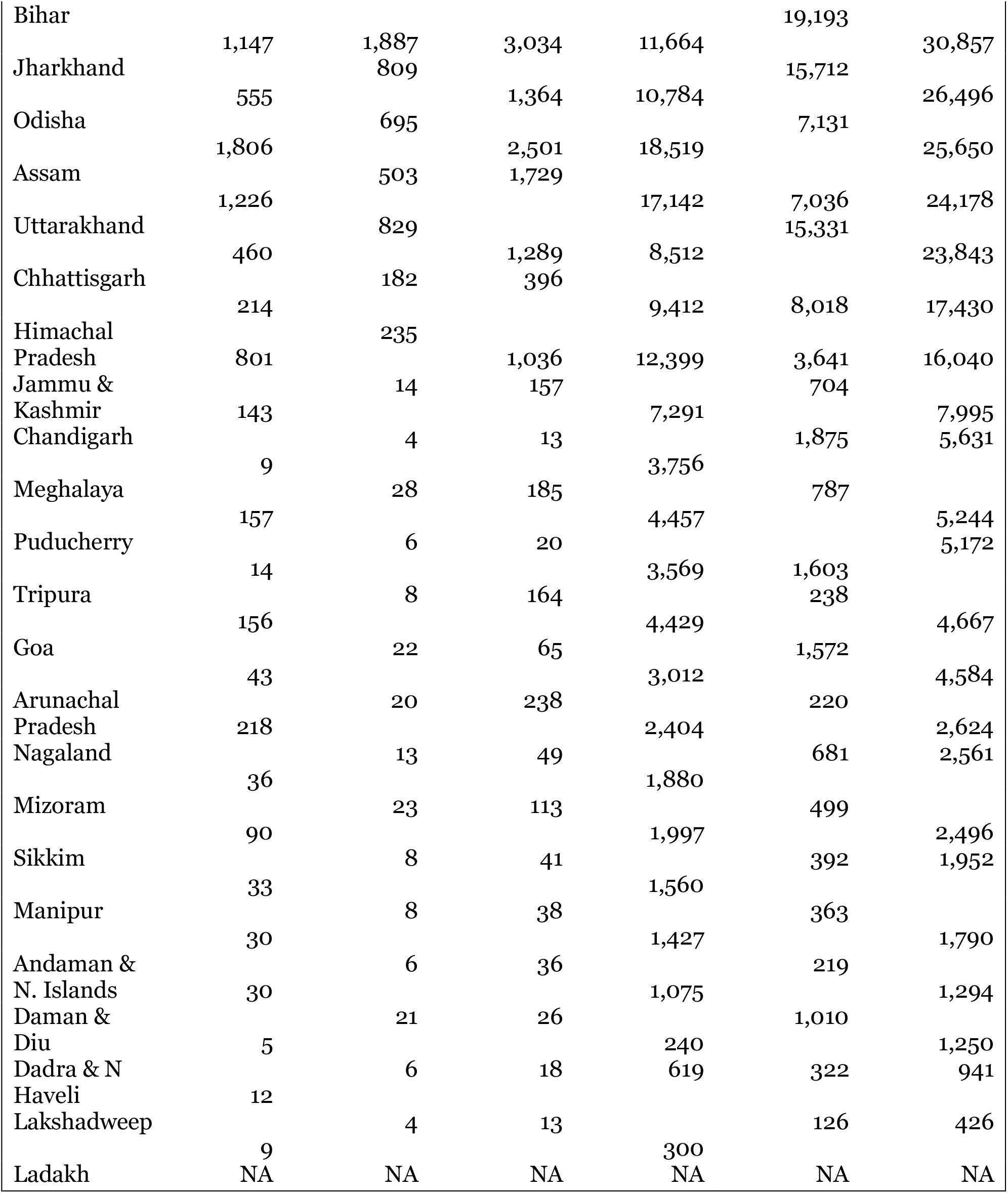
Hospitals and hospital beds in India, in descending order of total hospital beds

**Supplemental Table 2:**
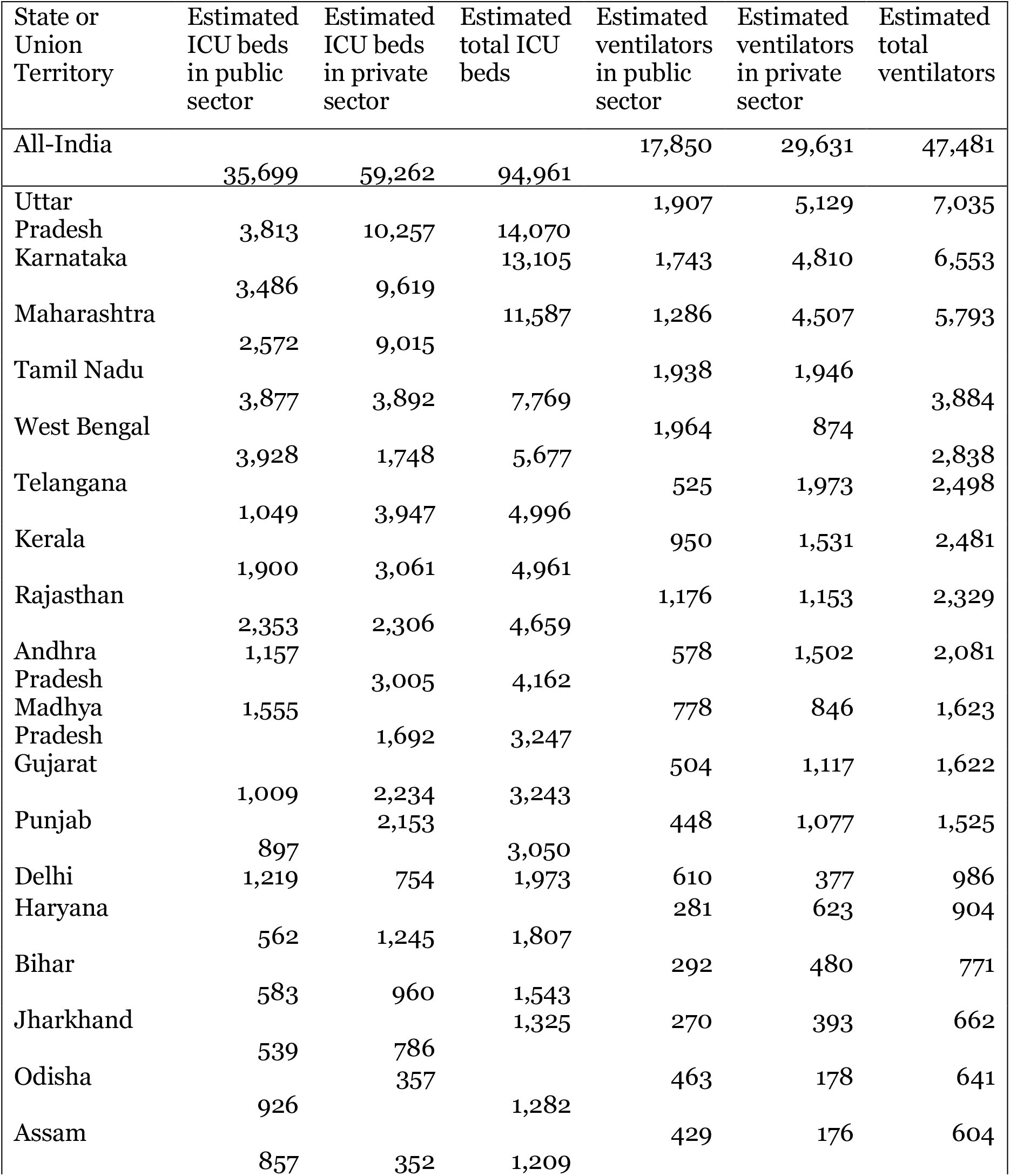
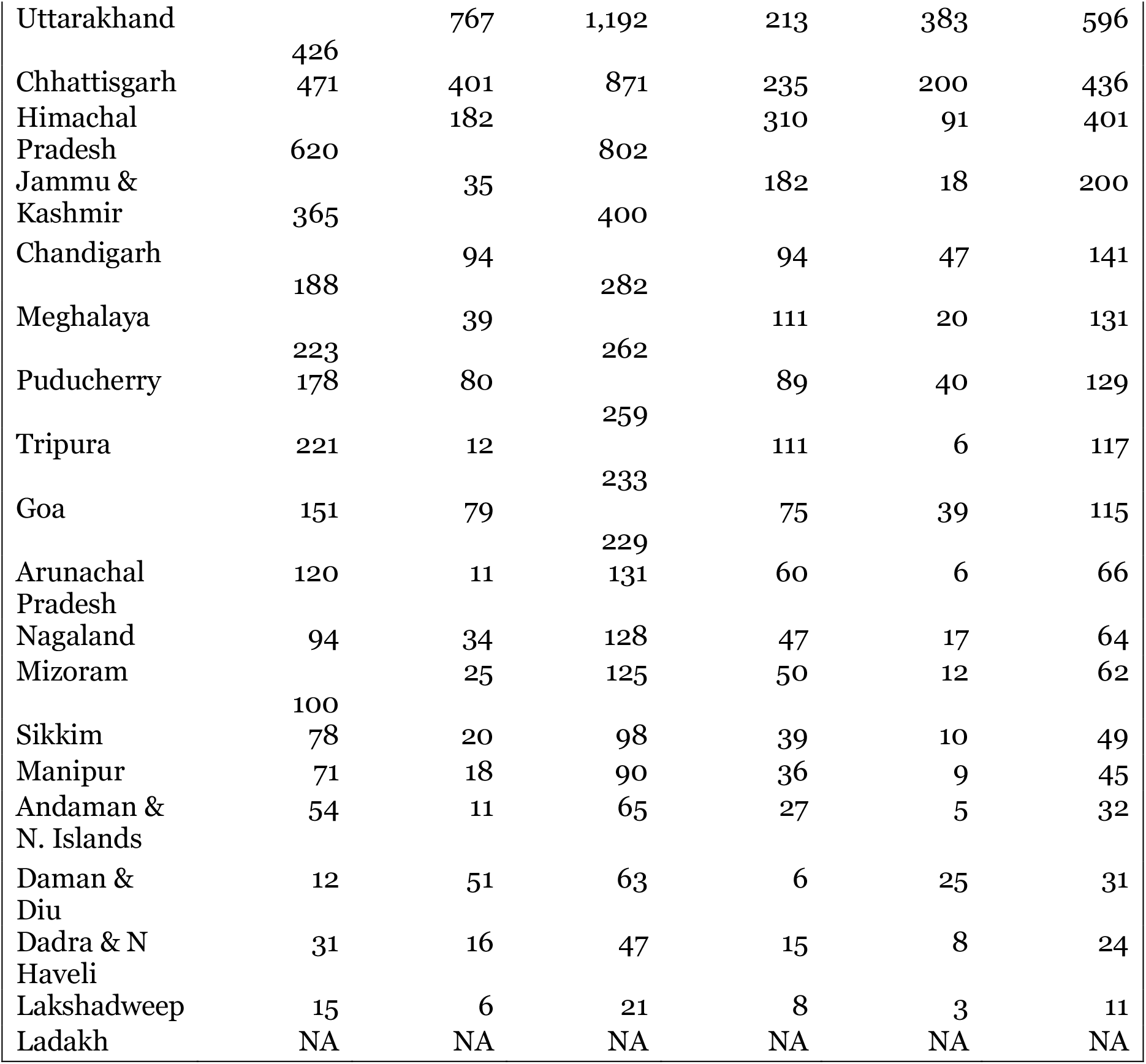
Estimated ICU beds and ventilators in India, in descending order of total ICU beds

**Supplemental Table 3:** Hospital beds per 100,000 in India, in descending order of total hospital beds

| State | Public Hospital<br>Beds per<br>100,000<br>Population | Private Hospital<br>Beds per<br>100,000<br>Population | Total Hospital<br>Beds per<br>100,000<br>Population |
| --- | --- | --- | --- |
| All India | 51.74 | 85.89 | 137.62 |
| Daman & Diu | 173.91 | 731.88 | 905.79 |
| Chandigarh | 340.22 | 169.85 | 510.07 |
| Karnataka | 104.17 | 287.45 | 391.62 |
| Puducherry | 258.62 | 116.19 | 374.82 |
| Sikkim | 282.61 | 71.09 | 353.70 |
| Andaman & Nicobar<br>Island | 259.66 | 52.81 | 312.47 |
| Lakshadweep | 217.39 | 91.40 | 308.79 |
| Goa | 181.88 | 94.96 | 276.84 |
| Kerala | 96.97 | 156.21 | 253.18 |
| Arunachal Pradesh | 217.75 | 19.97 | 237.72 |
| Telangana | 49.69 | 186.93 | 236.62 |
| Dadara & Nagar Havelli | 149.52 | 77.71 | 227.23 |
| Mizoram | 180.89 | 45.22 | 226.11 |
| Himachal Pradesh | 166.38 | 48.86 | 215.24 |
| Uttarakhand | 75.22 | 135.48 | 210.70 |
| Punjab | 60.16 | 144.47 | 204.63 |
| NCT of Delhi | 120.20 | 74.30 | 194.49 |
| Tamil Nadu | 87.24 | 87.59 | 174.83 |
| Maharashtra | 38.39 | 134.55 | 172.94 |
| Meghalaya | 140.42 | 24.78 | 165.20 |
| Nagaland | 113.53 | 41.14 | 154.67 |
| Andhra Pradesh | 40.40 | 104.93 | 145.33 |
| Uttar Pradesh | 35.20 | 94.68 | 129.88 |
| Rajasthan | 62.22 | 60.99 | 123.21 |
| Tripura | 110.67 | 5.95 | 116.62 |
| Haryana | 36.04 | 79.84 | 115.88 |
| West Bengal | 75.81 | 33.74 | 109.55 |
| Gujarat | 30.58 | 67.75 | 98.33 |
| Assam | 51.97 | 21.33 | 73.31 |
| Madhya Pradesh | 34.89 | 37.95 | 72.84 |
| Manipur | 57.45 | 14.63 | 72.08 |
| Jharkhand | 29.27 | 42.64 | 71.91 |
| Jammu & Kashmir | 56.21 | 5.42 | 61.63 |
| Chhattisgarh | 30.05 | 25.59 | 55.64 |
| Odisha | 39.47 | 15.20 | 54.67 |
| Bihar | 9.66 | 15.89 | 25.55 |

**Supplemental Table 4:** Estimated ICU beds and ventilators per capita in India, in descending order of total ICU beds

| State | Public ICU<br>Beds Per<br>100,000 | Private<br>ICU Beds<br>per<br>100,000 | Total ICU<br>Beds per<br>100,000 | Public<br>Ventilators<br>per<br>100,000 | Private<br>Ventilators<br>per<br>100,000 | Total<br>Ventilators<br>per<br>100,000 |
| --- | --- | --- | --- | --- | --- | --- |
| All India | 2.59 | 4.29 | 6.88 | 1.29 | 2.15 | 3.44 |
| Daman & Diu | 8.70 | 36.96 | 45.65 | 4.35 | 18.12 | 22.46 |
| Chandigarh | 17.03 | 8.51 | 25.54 | 8.51 | 4.26 | 12.77 |
| Karnataka | 5.21 | 14.37 | 19.58 | 2.60 | 7.19 | 9.79 |
| Puducherry | 12.90 | 5.80 | 18.70 | 6.45 | 2.90 | 9.35 |
| Sikkim | 14.13 | 3.62 | 17.75 | 7.07 | 1.81 | 8.88 |
| Andaman &<br>Nicobar Island | 13.04 | 2.66 | 15.70 | 6.52 | 1.21 | 7.73 |
| Lakshadweep | 10.87 | 4.35 | 15.22 | 5.80 | 2.17 | 7.97 |
| Goa | 9.12 | 4.77 | 13.89 | 4.53 | 2.36 | 6.88 |
| Kerala | 4.85 | 7.81 | 12.66 | 2.42 | 3.91 | 6.33 |
| Arunachal Pradesh | 10.87 | 1.00 | 11.87 | 5.43 | 0.54 | 5.98 |
| Telangana | 2.48 | 9.35 | 11.83 | 1.24 | 4.67 | 5.92 |
| Dadara & Nagar<br>Haveli | 7.49 | 3.86 | 11.35 | 3.62 | 1.93 | 5.56 |
| Mizoram | 9.06 | 2.26 | 11.32 | 4.53 | 1.09 | 5.62 |
| Himachal Pradesh | 8.32 | 2.44 | 10.76 | 4.16 | 1.22 | 5.38 |
| Uttarakhand | 3.76 | 6.78 | 10.54 | 1.88 | 3.38 | 5.27 |
| Punjab | 3.01 | 7.22 | 10.23 | 1.50 | 3.61 | 5.12 |
| NCT of Delhi | 6.01 | 3.72 | 9.73 | 3.01 | 1.86 | 4.87 |
| Tamil Nadu | 4.36 | 4.38 | 8.74 | 2.18 | 2.19 | 4.37 |
| Maharashtra | 1.92 | 6.73 | 8.65 | 0.96 | 3.36 | 4.32 |
| Meghalaya | 7.03 | 1.23 | 8.25 | 3.50 | 0.63 | 4.13 |
| Nagaland | 5.68 | 2.05 | 7.73 | 2.84 | 1.03 | 3.86 |
| Andhra Pradesh | 2.02 | 5.25 | 7.27 | 1.01 | 2.62 | 3.63 |
| Uttar Pradesh | 1.76 | 4.73 | 6.49 | 0.88 | 2.37 | 3.25 |
| Rajasthan | 3.11 | 3.05 | 6.16 | 1.56 | 1.52 | 3.08 |
| Tripura | 5.52 | 0.30 | 5.82 | 2.77 | 0.15 | 2.92 |
| Haryana | 1.80 | 3.99 | 5.79 | 0.90 | 2.00 | 2.90 |
| West Bengal | 3.79 | 1.69 | 5.48 | 1.90 | 0.84 | 2.74 |
| Gujarat | 1.53 | 3.39 | 4.92 | 0.76 | 1.69 | 2.46 |
| Assam | 2.60 | 1.07 | 3.67 | 1.30 | 0.53 | 1.83 |
| Madhya Pradesh | 1.74 | 1.90 | 3.64 | 0.87 | 0.95 | 1.82 |
| Jharkhand | 1.46 | 2.13 | 3.60 | 0.73 | 1.07 | 1.80 |
| Manipur | 2.86 | 0.72 | 3.58 | 1.45 | 0.36 | 1.81 |
| Jammu & Kashmir | 2.81 | 0.27 | 3.08 | 1.40 | 0.14 | 1.54 |
| Chhattisgarh | 1.50 | 1.28 | 2.78 | 0.75 | 0.64 | 1.39 |
| Odisha | 1.97 | 0.76 | 2.73 | 0.99 | 0.38 | 1.37 |
| Bihar | 0.48 | 0.80 | 1.28 | 0.24 | 0.40 | 0.64 |

